# Omitting biopsy in PI-RADS 3? The role of Stockholm3 and PSA density-findings from a prospective Swiss non-referral cohort

**DOI:** 10.64898/2026.01.12.26343907

**Authors:** Nicolas Arnold, Jonas Mudlagk, Odile Minder, Julian Cornelius, Carlo Di Bona, Beat Roth, Livio Mordasini, Nicola Giudici

## Abstract

**Introduction:** PI-RADS 3 (‘equivocal’) prostate MRI lesions pose a diagnostic challenge, particularly in non-referral settings with variable MRI quality. We evaluated whether prostate-specific antigen density (PSAD) and the Stockholm3 blood test can aid biopsy decisions for men with PI-RADS 3 lesions in a prospective, non-referral Swiss cohort.

**Methods:** Retrospective analysis of a prospective registry at a non-referral urology practice in Switzerland. From 2023–2025, men with PSA >1.5 ng/mL or suspicious digital rectal examination underwent Stockholm3 testing, MRI, and MRI-fusion plus systematic prostate biopsy. Clinically significant prostate cancer (csPCa) was defined as ISUP grade ≥2. Diagnostic accuracy and decision curve analysis compared Stockholm3 (≥13%, ≥15%, ≥18%) and PSAD thresholds (0.10, 0.15, 0.20ng/mL/cc) within the PI-RADS 3 subgroup.

**Results:** Overall, 50/174 (29%) men had PI-RADS 3 lesions, 59/174 (34%) were diagnosed with csPCa. Among PI-RADS 3 lesions, csPCa prevalence was 14%. Outcome distributions shifted towards higher ISUP grades with increasing PSAD thresholds when compared to baseline (≥0.15ng/mL/cc: 44% ISUP≥2; p=0.009), whereas Stockholm3 cut-offs showed no significant change for csPCa. As continuous measures, PSAD demonstrated superior discrimination (AUC 0.751; 95% CI 0.46-0.95) than Stockholm3 (AUC 0.595; 95% CI 0.30–0.88), confirming higher overall diagnostic accuracy within the PI-RADS 3 subgroup. Decision curve analysis demonstrated higher net benefit for PSAD-based approaches.

**Conclusion:** In non-referral practice, Stockholm3 did not reliably exclude csPCa among men with PI-RADS 3 lesions. PSAD-based triage, particularly at ≥0.15ng/mL/cc, showed superior diagnostic utility and may support safe biopsy deferral in selected cases. Prospective validation in broader non-referral settings is warranted.

## Introduction

Magnetic resonance imaging (MRI) with PI-RADS (Prostate Imaging Reporting and Data System) reporting has reshaped the prostate cancer diagnostic pathway [1]; yet PI-RADS 3 (‘equivocal’) lesions remain a gray zone. Meta-analyses estimate PI-RADS 3 in ∼17% of MRIs, with a clinically significant cancer yield of around 18–19% [2]. Performance varies by reader: low-volume/less-experienced radiologists show worse accuracy and tend to assign PI-RADS 3 more often [3–6]. Lower reliability of MRI further increases the risk of variation in prostate cancer treatment practice across non-referral institutions [7].

PSA density (PSAD) is a simple adjunct—readily available when MRI and PSA are known—that enhances risk stratification in this setting. Contemporary evidence and guidelines support risk-adapted strategies that combine PSAD cutoffs with MRI findings to refine patient selection and guide biopsy decisions [8–10].

The blood-based Stockholm3 test has been proposed as an improved diagnostic alternative, integrating clinical variables, blood biomarkers, and genetic markers into a single risk score [11]. Its effectiveness has been validated across different populations and countries, showing accuracy and reduction of unnecessary MRI and prostate biopsies[12–14]. Moreover, it has also been investigated in combination with MRI in several studies and settings, further supporting its role in optimizing diagnostic pathways [15–18].

Given the clinical relevance—particularly in non-referral settings where equivocal and less reliable MRI findings are more frequent—and the uncertainty surrounding PSAD and Stockholm3 cut-points, we investigated a prospective Swiss non-referral cohort to test the hypothesis that biopsy can be safely omitted in men with PI-RADS 3 lesions and Stockholm3 <15%, and to benchmark Stockholm3 against PSAD thresholds (0.10, 0.15, 0.20 ng/mL/cc). To our knowledge, this represents the first study to specifically address this question in a non-referral cohort.

## Methods

### Patient population and study design

Initial screening was performed in men with PSA >1.5 ng/mL or a suspicious digital rectal examination between November 2023 and January 2025 at a non-referral urology practice with multiple sites in Switzerland (Uroviva, Sursee, Switzerland). A total of 612 patients were prospectively included in the registry. For this retrospective analysis, we extracted data from 174 patients who underwent Stockholm3 testing, MRI and MRI-fusion plus systematic prostate biopsy. Men with a prior diagnosis of prostate cancer were excluded. Clinical, imaging, and histopathological data were collected, including age, family history, digital rectal examination findings, 5-ARI use, prior biopsy history, prostate volume, PSA, PSAD, Stockholm3 score, MRI findings with PI-RADS score, and biopsy results. The study was conducted in accordance with ethical standards laid down in the 1964 Declaration of Helsinki and its later amendments. All patients provided their consent by signing the Swiss General Consent form.

### Outcome Definition

The primary outcome was clinically significant prostate cancer (csPCa), defined as ISUP (International Society of Urological Pathology) grade ≥ 2 on biopsy. The secondary outcome comprised distribution of ISUP grade groups among patients with PI-RADS 3 according to different diagnostic PSAD and Stockholm3 thresholds, as well as the diagnostic performance of PSAD and Stockholm3 when evaluated both threshold-based and as continuous variables.

### Imaging and biopsy procedures

Multiparametric prostate MRI was performed at collaborating radiology centers and reported according to PI-RADS v2.1 by board-certified radiologists—who were not necessarily subspecialized in prostate imaging—across 21 centers. Biopsies were performed transrectally under local anesthesia and antibiotic prophylaxis. For patients with PI-RADS ≥ 3, three MRI-targeted cores per lesion were obtained in addition to a 12-core systematic template biopsy. Histopathology was reported using ISUP grading.

### Index tests and comparators

Stockholm3 test analysis, requiring a 12ml blood sample, was performed by accredited laboratories (A3P Biomedical, Uppsala, Sweden, or Labor Team W AG, Goldach, Switzerland). The test integrates clinical variables (age, family history of prostate cancer, previous biopsy), plasma protein biomarkers (total PSA, free PSA, intact PSA, hK2, MSMB, MIC1), and > 100 genetic polymorphisms into a proprietary algorithm, and is reported as a continuous risk percentage of detecting csPCa (ISUP Grade ≥ 2). We evaluated cut-offs of ≥ 13%, ≥ 15%, and ≥ 18% and as a continuous variable. PSAD was calculated as PSA (ng/mL) divided by prostate volume (mL) and analyzed both as a continuous variable and at thresholds of ≥ 0.10, ≥ 0.15, and ≥ 0.20 ng/mL/cc.

### Statistical analysis

Basic patient characteristics were analyzed using descriptive statistics. Continuous variables are presented as medians with interquartile ranges (IQR), and categorical variables as counts and percentages. Analyses then focused on men with PI-RADS 3 lesions. For each decision rule, PSAD thresholds and Stockholm3 cut-offs, we calculated sensitivity, specificity, positve predictive value (PPV), and negative predictive value (NPV), each with 95% confidence intervals, using csPCa (ISUP ≥ 2) at biopsy as outcome. Outcome distributions under each strategy were compared with the baseline PI-RADS 3 distribution using chi-square tests, with Fisher’s exact test applied when appropriate. The distribution ISUP ≥ 3 categories was reported descriptively. We performed decision curve analysis within the PI-RADS 3 subgroup, comparing PSAD- and Stockholm3-based strategies with biopsy-all and biopsy-none across a range of clinically plausible threshold probabilities, with net benefit as the outcome metric (endpoint: ISUP ≥ 2). All analyses were prespecified and performed in R (v4.5.1), with p <0.05 considered statistically significant.

## Results

### Patient characteristics

A total of 174 patients were included. Median age was 65.0 years (IQR 60.0–70.0). Eighteen (10%) had a prior negative biopsy, and 30 (19%) reported a positive family history for prostate cancer. Median PSA was 5.3 ng/mL (IQR 4.4–7.1), and the median Stockholm3 score was 17 (IQR 13.0–25.0). Digital rectal examination was performed in 74 (43%) men, of whom 5/74 (7%) were abnormal. The median prostate volume was 53.5 mL (IQR 40.0–71.0). Regarding MRI findings, 92 patients (53%) had PI-RADS 4–5, 50 (29%) had PI-RADS 3, and 32 (18%) had PI-RADS 1–2. While biopsy outcomes were benign in 91 (52%), ISUP grade 1 was present in 24 cases (14%). Fifty-nine patients (34%) had clinically significant cancer (ISUP ≥ 2): 31 (18%) grade 2, 16 (9%) grade 3, 8 (5%) grade 4, and 4 (2%) grade 5 (Table 1).

**Table 1.**
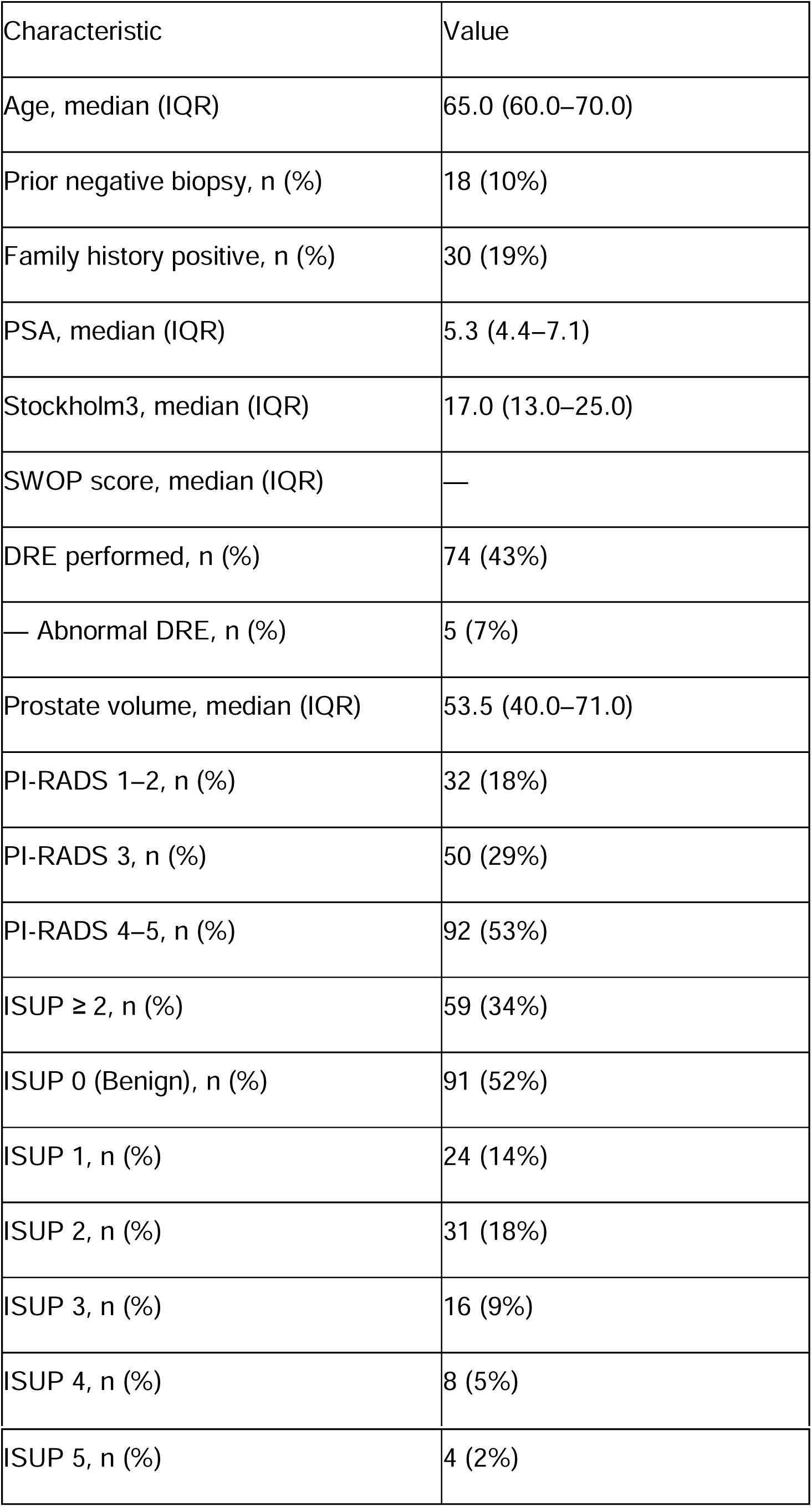
Patients Characteristics.

| Characteristic | Value |
| --- | --- |
| Age, median (IQR) | 65.0 (60.0–70.0) |
| Prior negative biopsy, n (%) | 18 (10%) |
| Family history positive, n (%) | 30 (19%) |
| PSA, median (IQR) | 5.3 (4.4–7.1) |
| Stockholm3, median (IQR) | 17.0 (13.0–25.0) |
| SWOP score, median (IQR) | — |
| DRE performed, n (%) | 74 (43%) |
| — Abnormal DRE, n (%) | 5 (7%) |
| Prostate volume, median (IQR) | 53.5 (40.0–71.0) |
| PI-RADS 1–2, n (%) | 32 (18%) |
| PI-RADS 3, n (%) | 50 (29%) |
| PI-RADS 4–5, n (%) | 92 (53%) |
| ISUP $\geq 2$, n (%) | 59 (34%) |
| ISUP 0 (Benign), n (%) | 91 (52%) |
| ISUP 1, n (%) | 24 (14%) |
| ISUP 2, n (%) | 31 (18%) |
| ISUP 3, n (%) | 16 (9%) |
| ISUP 4, n (%) | 8 (5%) |
| ISUP 5, n (%) | 4 (2%) |

### Outcome distribution in PI-RADS 3 by strategy

In the PI-RADS 3 subgroup (n = 50), the baseline outcome mix was 70% benign, 16% ISUP 1, 14% ISUP ≥ 2, with 4% ISUP ≥ 3. Applying a PSAD threshold ≥ 0.10 ng/mL/cc selectively increased the proportion of higher-grade disease within this subgroup (27% ISUP ≥ 2; p = 0.041). The PSAD ≥ 0.15 ng/mL/cc threshold further enriched for csPCa (44% ISUP ≥ 2; p = 0.009), whereas PSAD ≥ 0.20 ng/mL/cc identified fewer patients (33% ISUP ≥ 2) and did not differ significantly from baseline (p = 0.232). In contrast, none of the Stockholm3 cut-offs (≥ 13%, ≥ 15%, ≥ 18%) significantly altered the outcome distribution compared with the baseline PI-RADS 3 group (p ≥ 0.28), with ISUP ≥ 2 rates ranging from 12% to 22% across thresholds (Table 2).

**Table 2.** Outcome distribution by strategy (PI-RADS 3 base) P values indicate comparison of ISUP grade distribution for each decision rule versus the overall PI-RADS 3 baseline group (chi-square or Fisher’s exact test).

| Strategy | Benign n (%) | ISUP 1 n (%) | ISUP ≥ 2 n (%) | ISUP ≥ 3 n (%) | P value |
| --- | --- | --- | --- | --- | --- |
| All | 35 (70%) | 8 (16%) | 7 (14%) | 2 (4%) | — |
| PSAD ≥ 0.10 | 12 (55%) | 4 (18%) | 6 (27%) | 2 (9%) | 0.041 |
| PSAD ≥ 0.15 | 5 (56%) | 0 (0%) | 4 (44%) | 1 (11%) | 0.009 |
| PSAD ≥ 0.20 | 4 (67%) | 0 (0%) | 2 (33%) | 0 (0%) | 0.232 |
| Stockholm3 ≥ 13% | 27 (68%) | 8 (20%) | 5 (12%) | 2 (5%) | 0.287 |
| Stockholm3 ≥ 15% | 18 (67%) | 5 (19%) | 4 (15%) | 2 (7%) | 0.838 |
| Stockholm3 ≥ 18% | 12 (67%) | 2 (11%) | 4 (22%) | 2 (11%) | 0.402 |

### Diagnostic performance in PI-RADS 3: PSAD and Stockholm3 thresholds

In the PI-RADS 3 subgroup (n = 50), PSAD ≥ 0.10 ng/mL/cc achieved a sensitivity of 85.7% (95% CI 48.7–97.4) and NPV 96.4% (95% CI 82.3–99.4) at a specificity of 62.8% (95% CI 47.9–75.6). Increasing the threshold to PSAD ≥ 0.15 improved specificity to 88.4% (95% CI 75.5–94.9) while maintaining moderate sensitivity (57.1%, 95% CI 25.0–84.2), and PSAD ≥ 0.20 further maximized specificity (90.7%, 95% CI 78.4–96.3) but with reduced sensitivity (28.6%, 95% CI 8.2–64.1).

For the Stockholm3 test, diagnostic performance remained modest across thresholds: ≥ 13% yielded sensitivity of 71.4% (95% CI 35.9–91.8) and specificity of 18.6% (95% CI 9.7–32.6); ≥ 15% achieved sensitivity 57.1% (95% CI 25.0–84.2) and specificity 46.5% (95% CI 32.5–61.1); and ≥ 18% improved specificity to 67.4% (95% CI 52.5–79.5) with unchanged sensitivity (57.1%, 95% CI 25.0–84.2). Corresponding PPV/NPV values ranged from 12.5–22.2% and 80.0–90.6%, respectively. When analyzed as continuous variables, PSAD demonstrated superior discrimination with an area under the receiver operating characteristic curve (AUC) of 0.751 (95% CI 0.46–0.95) compared with Stockholm3 (AUC 0.595, 95% CI 0.30–0.88), confirming higher overall diagnostic accuracy of PSAD within the PI-RADS 3 subgroup (Table 3).

**Table 3.** Performance of PSAD. (≥ 0.10/0.15/0.20) and Stockholm3 (≥ 15%) to detect ISUP ≥ 2 in PI-RADS 3

| Rule | Sensitivity (95% CI) | Specificity (95% CI) | PPV (95% CI) | NPV (95% CI) | AUC (95% CI)* |
| --- | --- | --- | --- | --- | --- |
| PSAD $\geq 0.10$ | 85.7 (48.7–97.4) | 62.8 (47.9–75.6) | 27.3 (13.2–48.2) | 96.4 (82.3–99.4) | 0.751 (0.46–0.95) |
| PSAD $\geq 0.15$ | 57.1 (25.0–84.2) | 88.4 (75.5–94.9) | 44.4 (18.9–73.3) | 92.7 (80.6–97.5) | |
| PSAD $\geq 0.20$ | 28.6 (8.2–64.1) | 90.7 (78.4–96.3) | 33.3 (9.7–70.0) | 88.6 (76.0–95.0) | |
| Stockholm3 $\geq 13\%$ | 71.4 (35.9–91.8) | 18.6 (9.7–32.6) | 12.5 (5.5–26.1) | 80.0 (49.0–94.3) | 0.595 (0.30–0.88) |
| Stockholm3 $\geq 15\%$ | 57.1 (25.0–84.2) | 46.5 (32.5–61.1) | 14.8 (5.9–32.5) | 87.0 (67.9–95.5) | |
| Stockholm3 $\geq 18\%$ | 57.1 (25.0–84.2) | 67.4 (52.5–79.5) | 22.2 (9.0–45.2) | 90.6 (75.8–96.8) | |

### Decision curve analysis

Figure 1 illustrates the decision curve analysis comparing diagnostic strategies for biopsy decision-making in men with PI-RADS 3 lesions: biopsy-all, biopsy-none, PSAD-based thresholds, and Stockholm3 cut-offs. Across clinically relevant threshold probabilities, PSAD-based strategies consistently provided higher net benefit than both default approaches. Among them, PSAD ≥ 0.15 ng/mL/cc demonstrated the most balanced performance, yielding clinical benefit at lower thresholds and maintaining discrimination up to approximately 0.45. In contrast, Stockholm3-based strategies—regardless of the selected cut-off—offered minimal or no additional net benefit compared with biopsy-all or biopsy-none, confirming the superior clinical utility of PSAD in this setting.

**Figure 1.**
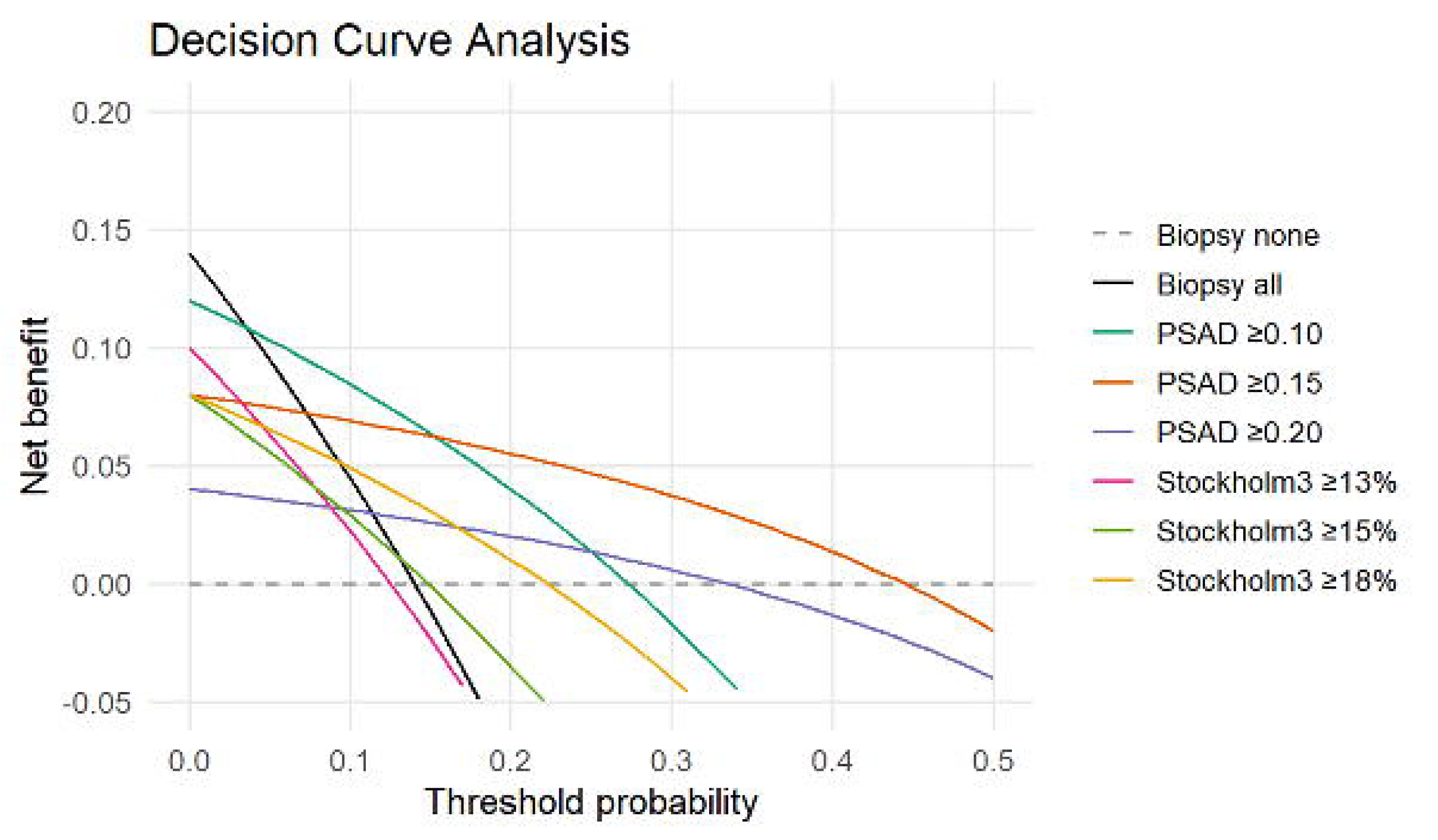
Decision curve analysis for four diagnostic strategies in men with PI-RADS 3 lesions.

## Discussion

In this prospective non-referral Swiss cohort of men with PI-RADS 3 lesions, the prevalence of csPCa (ISUP ≥ 2) was 14%. Across Stockholm3 thresholds, csPCa detection remained largely unchanged, ranging from 12.5% at ≥ 13% to 22.2% at ≥ 18%, with no significant difference compared to baseline. In contrast, PSAD demonstrated a clear, stepwise association with csPCa risk—rising from 3.6% at <0.10 ng/mL/cc to 15.4% at 0.10–0.15, 66.7% at 0.15–0.20, and 33.3% at ≥ 0.20 ng/mL/cc. Diagnostic accuracy analyses confirmed superior overall discrimination for PSAD (AUC 0.751, 95% CI 0.46–0.95) compared with Stockholm3 (AUC 0.595, 95% CI 0.30–0.88), consistent across all tested cut-offs (≥ 13%, ≥ 15%, ≥ 18%), highlighting PSAD’s greater ability to distinguish clinically significant disease in the PI-RADS 3 subgroup. Decision-curve analysis demonstrated that PSAD-based strategies provided consistently higher net benefit than biopsy-all or biopsy-none approaches, and clearly outperformed all Stockholm3-based rules across clinically relevant thresholds.

Our findings complement and extend existing evidence on risk stratification for men with PI-RADS 3 lesions [8, 9]. In our cohort, the proportion of PI-RADS 3 lesions (29%) was higher than typically reported in referral or expert-reader series, where PI-RADS 3 accounts for approximately 10–17% of prostate MRIs [9, 19, 20]. This likely reflects the greater reader variability and lower MRI specialization in non-referral practice [3–7], supporting the need for additional objective tools to guide biopsy decisions in this context. Different studies provide consistent quantitative support for PSAD in the PI-RADS 3 setting [21–23]. Drevik et al. demonstrated a sensitivity of 86.4% for the detection of csPCa when applying a PSAD threshold of ≥ 0.15 ng/mL/cc to guide biopsy decisions in patients with PI-RADS 3 lesions [21]. Nguyen et al. found that a PSAD cutoff of 0.15 ng/mL/cc achieved a sensitivity of 72% and specificity of 79% for detecting csPCa, offering the best diagnostic balance while cautioning against its isolated use [22]. Fang et al. reported similar findings in a multi-institutional cohort, where PSAD ≥ 0.15 ng/mL/cc yielded an AUC of 0.74 and remained independently predictive of csPCa, with stronger performance in biopsy-naïve men [23]. Furthermore, Pellegrino et al. investigated PSAD thresholds in men with negative MRI (PI-RADS ≤ 2) and found that the commonly used cutoff of 0.15 ng/mL/cc is justified only under conditions of low MRI accuracy, recommending a higher threshold of ≥ 0.20 for average or high-quality MRI [9].

Our results align with these observations: PSAD ≥ 0.15 ng/mL/cc identified a subgroup enriched for csPCa while maintaining high specificity, and decision-curve analysis indicated superior net benefit compared with biopsy-all or Stockholm3-based strategies. By contrast, the Stockholm3 test—though previously shown to reduce unnecessary biopsies in PSA-based pathways and combined MRI workflows—performed less effectively in our PI-RADS 3 subgroup [16]. Prior validations largely stem from screening or referral cohorts with centralized high-quality MRI reading whereas our study represents, to our knowledge, the first evaluation in a real-world, non-referral setting with heterogeneous MRI quality and reporting. In such environments, PI-RADS 3 lesions likely include a mix of true low-risk and misclassified higher-risk cases, highlighting the need for discriminatory tools in this context. In contrast, PSAD—a simple biologically grounded measure reflecting PSA production relative to gland volume—appears more robust across settings.

In non-referral settings, where MRI interpretation tends to be more variable and of lower quality, PSAD-based triage offers a pragmatic and cost-neutral approach to guide biopsy decisions in men with PI-RADS 3 lesions. Applying a PSAD threshold of <0.10 ng/mL/cc can help identify a subgroup at very low risk (3.6% csPCa in our cohort) for whom surveillance rather than immediate biopsy may be appropriate. In contrast, our findings indicate that Stockholm3 alone cannot reliably identify such low-risk men in this context. Because PSAD is readily available, easily calculated, and cost-neutral, it can be seamlessly integrated into everyday clinical practice to support more selective and evidence-based biopsy decisions without requiring additional infrastructure. Future multicenter studies should refine integrated models combining both metrics while accounting for local imaging quality and experience.

Several limitations should be acknowledged. First, the diagnostic algorithm was not standardized, as the decision to use the Stockholm3 test was left to the discretion of the treating physician. However, it also reflects real-world practice in non-referral settings, where testing decisions are influenced by individual clinical judgment, patient preferences, and local healthcare economic factors [24]. Second, although our registry was prospective, the analysis is retrospective, which may introduce selection bias and unmeasured confounding. Third, MRI acquisitions and reporting occurred across 21 centres with radiologists not all subspecialised in prostate imaging, and pathology was not centrally reviewed—while this reduces internal standardization, it enhances real-world generalisability to non-referral settings where most Stockholm3/MRI validation studies are done. Fourth, the number of men with PI-RADS 3 lesions and csPCa was relatively small, limiting precision and widening confidence intervals; thus, findings are exploratory. Fifth, we applied predefined

Stockholm3 thresholds which may not be optimal in this specific context. Moreover, long-term prostatectomy pathology and clinical outcome data were not available, limiting the assessment of disease progression and potential treatment impact. Given these limitations, prospective multicentre studies in non-referral settings are needed to confirm our observations.

Although the study has several limitations, its real-world design represents a major strength. The interpretation of prostate MRIs was performed across various radiology practices by radiologist not necessarily subspecialized in prostate imaging, reflecting routine clinical procedures rather than idealized study conditions. Furthermore, the clear structure and standardized workflow mirror everyday practice and thus most most real-world clinical settings. Therefore, the study provides valuable and highly relevant insights into current clinical practice and enhances the generalisability of its findings.

## Conclusion

In this prospective, non-referral cohort of men with PI-RADS 3 lesions in the primary diagnostic setting, the Stockholm3 test alone did not demonstrate adequate rule-out performance, resulting in a non-negligible rate of missed clinically significant prostate cancers. Therefore, biopsy should not be omitted on the basis of Stockholm3 alone when applied in real-world non-referral settings. PSAD-based triage may improve diagnostic safety; however, threshold selection should be individualized and integrated within a shared decision-making framework.

## Data Availability

The data underlying this article will be shared on reasonable request to the corresponding author.

